# The impact of bilateral versus unilateral anterior temporal lobe damage on face recognition, person knowledge and semantic memory

**DOI:** 10.1101/2024.02.10.24302526

**Authors:** Matthew A. Rouse, Siddharth Ramanan, Ajay D. Halai, Angélique Volfart, Peter Garrard, Karalyn Patterson, James B. Rowe, Matthew A. Lambon Ralph

## Abstract

In recent years, the functional importance of the anterior temporal lobes (ATLs) has come to prominence in two active, albeit unconnected branches of the literature. In one branch, neuropsychology and functional neuroimaging evidence emphasises the role of the ATLs in face recognition and linking faces to biographical knowledge. In the other, cognitive and clinical neuroscience investigations have shown that the ATLs are critical to all forms of semantic memory. To draw these literatures together and generate a unified account of ATL function, we test the predictions arising from each literature and examine the effects of bilateral *versus* unilateral ATL damage on face recognition, person knowledge and semantic memory. Sixteen people with bilateral ATL atrophy from semantic dementia (SD), 17 people with unilateral ATL resection for temporal lobe epilepsy (TLE; left=10, right=7), and 14 controls completed a test battery encompassing general semantic processing, person knowledge and perceptual face matching. SD patients were severely impaired across all semantic tasks, including person knowledge. Despite commensurate total ATL damage, unilateral resection generated mild impairments, with minimal differences between left- and right-ATL resection. Face matching performance was largely preserved but slightly reduced in SD and right TLE. All groups displayed the classic familiarity effect in face matching; however, this benefit was reduced in SD and right TLE groups and was aligned with the level of item-specific semantic knowledge in all participants. We propose a unified neurocognitive framework whereby the ATLs underpin a resilient bilateral representation system that supports semantic memory, person knowledge and face recognition.

## Introduction

The role of the anterior temporal lobes (ATLs) has become of key interest to cognitive neuroscientists in recent years, resulting in two very active but largely distinct research pursuits. There is evidence from neuropsychology and functional neuroimaging that the ATLs are important for face recognition/person knowledge (1–4). A separate literature implicates the ATLs in multimodal semantic memory, including knowledge of familiar faces/people alongside all other concepts (5, 6). These two research areas and associated theories have remained largely separate from each other, despite making potentially complementary predictions. The aim of the current study was to bridge the two literatures - and generate a unified neurocognitive framework for the role of the ATLs in face recognition, person knowledge and semantic processing. Accordingly, a bespoke neuropsychological battery was used to assess the effect of bilateral vs. unilateral ATL damage on (i) general semantic memory vs. person knowledge and (ii) perceptual face matching of famous and unfamiliar faces. To identify the effects of bilateral vs. unilateral (left vs. right) ATL damage, we compared two patient groups associated with ATL damage; semantic dementia (SD, including semantic variant primary progressive aphasia and ‘right’ semantic dementia), characterised by bilateral ATL atrophy from neurodegeneration (7–10), and people who had undergone left or right unilateral ATL resection for temporal lobe epilepsy (TLE).

There is evidence from positron emission tomography (11–14), functional magnetic resonance imaging (fMRI) (15–18) and intracranial electrode recordings (19) that the ATLs respond to familiar faces, as well as neuropsychological demonstrations of impaired face recognition following ATL damage from neurodegenerative disorders or unilateral resection (20–25). Based on these findings, neurocognitive models of face recognition have broadened to include the ATL as part of an extended network critical for linking faces with stored semantic knowledge (1–4). Indeed, the existence of face-selective patches in the ATL has been proposed (4, 17), thought to be homologous to the anterior face patches identified in macaques (26–28).

In contrast to a face-specific function, there is convergent evidence from neuropsychology, fMRI, transcranial magnetic stimulation (TMS) and grid electrode studies that the ATLs are critical for supporting semantic memory more broadly (29–33). Perhaps most strikingly, people with SD, characterised by bilateral ATL atrophy, display a global degradation of conceptual knowledge (7–9, 29). This semantic degradation occurs for all types of concepts, including but not limited to knowledge of familiar faces/people (5, 6, 10, 29, 34). The ATLs have been considered to underpin a transmodal, transtemporal semantic hub, which supports the creation of generalisable concepts from the numerous multimodal experiences we have of each concept over our lifetimes (5, 6).

FMRI studies consistently detect strong bilateral ventrolateral ATL activation in relation to all types of conceptual knowledge (30), provided techniques are utilised to maximise ATL signal (35). A recent study found that this same region activated in response to both the faces and spoken names of famous people and to specific-level concepts other than people, such as famous landmarks. An anterior extension to this core vATL region demonstrated weaker yet more selective activation for people over the other categories (overlapping with the peaks described in the face recognition literature) (36).

ATL damage does not generate the perceptual face processing deficits associated with damage to posterior temporal cortex (e.g., the fusiform face area) (37, 38). People with SD perform at normal levels on tasks of perceptual matching of unfamiliar faces, which require distinguishing between faces but do not require activation of specific conceptual knowledge (20, 25). Healthy participants match famous/familiar faces faster and more accurately than unfamiliar faces (39–42), which has led some researchers to argue for qualitative differences in the perceptual processing of familiar and unfamiliar faces (39). One potentially important difference is that familiar faces are laden with specific semantic knowledge, which may support and facilitate face processing, whereas perception of unfamiliar faces can only rely on visual features (43, 44). Therefore, although not *critical*, the ATLs may enhance performance in tasks that require perceptual matching of faces through feedback activation, thus reducing perceptual demands and contributing to the known familiarity effect (43, 44). There is behavioural evidence for an interaction between conceptual and perceptual information in healthy participants, where associating previously unfamiliar stimuli with conceptual labels improves later recognition (45, 46), and from people with SD who are impaired on tasks which require successfully classifying visually different exemplars of objects as the ‘same thing’, or words/objects as real versus non-real (47–49).

The differential function of the left and right ATLs is a key area of debate (50). Face recognition theories have generally made no strong claims regarding left/right ATL differences (1, 3), however the early stages of face perception are thought to be supported bilaterally with a right-sided dominance (51–54). Neuropsychological studies have implicated the importance of the right ATL in face recognition, based on several case reports of face recognition problems after right ATL damage from either SD or unilateral resection (23, 24, 55, 56). The underlying explanation for this right ATL bias is debated, with suggestions that the right ATL is specialised for representing transmodal person-specific semantic knowledge, rather than faces specifically (57) or alternatively that the right ATL is specialised for retrieving semantic information from visual inputs (i.e., faces) whereas the left ATL is important for retrieving verbal semantics (e.g., written and spoken names) (10, 34). The evidence from functional neuroimaging is less clear cut, with evidence for bilateral ATL activation in response to faces or people’s names (36, 58).

A hub and spoke model of semantic memory has been proposed in which the bilateral ATLs work in concert to support transmodal semantic representations, and that bilateral neural implementation can make functional systems more resilient to unilateral damage (5, 59, 60). This framework does not deny emergent functional differences between the left and right ATLs but, in line with various computational modelling demonstrations, suggests that these differences could be a consequence of differential connectivity of the left/right ATL with modality-specific cortical regions (51, 61–63). The right posterior temporal cortex is more dominant for face processing (51, 64) and so consequently the face recognition problems associated with right ATL damage may be because the right ATL receives increased visual input from right posterior temporal areas (64).

To determine the impact of bilateral vs. unilateral damage and the relative contributions of the left/right ATL to semantics and face recognition, we directly compared people with semantic dementia to people with unilateral ATL resection, using the same neuropsychological and structural imaging measures. Although ATL abnormalities can be somewhat asymmetric in SD patients, especially initially, there is always hypometabolism and indeed some atrophy in the contralateral ATL (21, 65); and with progression of the disease, damage on the initially less affected side catches up (66). This bilateral damage in SD contrasts with unilateral ATL resection which provides insights about the individual contributions of the left and right ATL (67, 68).

## Materials and Methods

### Participants

Sixteen people with SD were recruited from specialist neurology clinics at Addenbrooke’s Hospital, Cambridge (*N*=11), John Radcliffe Hospital, Oxford (*N*=4) and St George’s Hospital, London *(N*=1). Seventeen people who had undergone unilateral en bloc anterior temporal lobectomy for TLE (left TLE=10, right TLE=7) were recruited from neuropsychology departments at Salford Royal NHS Foundation Trust, Manchester (*N*=8) and Walton Centre NHS Foundation Trust, Liverpool (*N*=9). All the ATL-resected cases had had late-onset, long-standing TLE stemming from unilateral hippocampal sclerosis, were left language dominant based on results of the Wada test and were in the chronic stage post-surgery. Normative data were obtained from fourteen healthy volunteers with no history of neurological or psychiatric disorders, recruited from the MRC Cognition and Brain Sciences Unit, University of Cambridge.

### General semantic memory

General semantic memory was assessed using a battery of receptive and expressive tasks, comprising the modified picture-version of Camel and Cactus semantic association test (mCCT) (69, 70), a synonym judgement task (71, 72), the Cambridge Naming (70, 72) and Boston Naming (72, 73) tests and a word-to-picture matching task. For all patients (*N*=33), a principal component analysis (PCA) was conducted on the mCCT, synonym judgement task and word-to-picture matching task scores. The PCA generated one component with an eigenvalue greater than one (2.69) which explained 89.5% of the total variance (Kaiser-Meyer-Olkin statistic =0.68). All three tasks loaded heavily onto this component (mCCT=0.92, synonym judgement task =0.94, word-to-picture matching task=0.98), and so factor scores were used as a composite score of total semantic impairment. The lower bound of normality for the composite score was derived by calculating the factor score of a hypothetical individual scoring 1.96 standard deviations below the control mean on all three tasks. Global cognitive function was assessed by the Addenbrooke’s Cognitive Examination-Revised (74) and executive functioning assessed using the Brixton Spatial Anticipation Test (75) and Raven’s Coloured Progressive Matrices B (76).

### Person knowledge

Two 44-item tasks were designed to assess person knowledge: face-to-name matching and face-to-profession matching. In each task, participants were shown a photograph of a famous person and instructed to point to the correct name/profession from four possible response options. Participants also completed a difficulty matched 42-item landmark-to-name matching task, to determine whether any person knowledge deficits were selective, or occurred for another type of specific-level concept. Performance accuracy on the person knowledge tasks was negatively correlated with age in controls (face-name matching; *r* = -0.63, *P* < 0.05, face-profession matching; *r* = -0.56, *P* < 0.05) and so groups were compared using Quade’s non-parametric ANCOVA (post-hoc Tukey HSD tests) with age included as a covariate. Individual patient deficits were determined using one-tailed modified t-tests. This method tests whether an individual’s score on a task is significantly below a control sample, and is recommended when comparing against small control samples (77). For the two person knowledge tasks, single case deficits were tested using Bayesian methods to control for the effect of age (78).

### Perceptual face matching

A face matching task (79) was administered on a laptop using E-prime software (version 1.2, Psychology Software Tools). Participants were presented with a triad of faces; one at the top of the screen and two below and were instructed to select which of the two faces below was the same person as the face at the top. Participants performed practice trials to ensure they understood the task, and accuracy and response times (RT) were recorded. There was no time limit, but participants were instructed to respond as quickly as they could. There were 44 items in total, where half of the faces were famous (using people from the person knowledge tasks as targets or foils) and half were unfamiliar. Each face was presented both upright and inverted on separate occasions. Only trials receiving correct responses were included in the RT analysis. Outlier RTs for each participant (1.96 standard deviations away from the participant’s mean RT) were replaced by their mean RT across all conditions (80). To assess the relationship between semantic knowledge and perception, each participant’s person knowledge results were used to categorise their face perception trials into ‘fully known’ (correct in both person knowledge tasks), ‘partially known’ (correct in only one person knowledge task) or ‘unknown’ (incorrect in both person knowledge tasks). Friedman’s ANOVA and post-hoc Wilcoxon tests (Holm-method for multiple comparisons) were conducted to compare RTs across different levels of semantic knowledge, as well as compared to RTs for unfamiliar face matching trials.

### Magnetic Resonance Imaging

Thirty participants (16 SD, 14 controls) completed a 3T structural MRI scan on a Siemens PRISMA at the MRC Cognition and Brain Sciences Unit or the Wolfson Brain Imaging Centre (both in Cambridge). The TLE group’s structural MRI scans on a 3T Phillips Achieva scanner were available from a previous study (68, 81). One TLE participant had undergone further ipsilateral temporal neurosurgery since his scan and so was excluded from the imaging analysis. MRI scans from 20 controls scanned for the original TLE study were included in the imaging analysis, so that TLE groups could be compared to a group matched for both age and scanning site (68, 81).

Voxel-based morphometry (VBM) was conducted to determine grey matter volumetric differences between patient groups and controls. A separate general linear model was created for each patient group versus controls, with age, intracranial volume, and scanning site included as covariates. An explicit mask was used which excluded any voxels for which >20% of participants had an intensity value of <0.1; this is a method recommended for analysis of atrophied brains (82). Significant clusters were extracted using a voxelwise statistical threshold of *P* < 0.05 (FWE-corrected) with a cluster threshold of 100 voxels. To visualise both the total amount as well as the distribution of ATL volume loss, grey matter intensities in the left and right ATL were extracted for each participant (using binarised masks derived from a previous ALE meta-analysis (58)). For each patient, values were z-scored relative to the control sample to calculate two indices; (i) ATL magnitude (sum of left and right ATL volume) and (ii) ATL asymmetry (left ATL volume minus right ATL volume) (21, 83, 84).

Since structural MRI is insensitive to some markers of neurodegeneration such as hypometabolism (65) and synaptic loss (85), VBM may underestimate ATL damage in SD. However, this caveat is relevant to unilateral resection cases too, where there may be additional damage secondary to the site of resection, such as white matter connectivity changes consistent with Wallerian degeneration (86, 87). The chronic epilepsy in the TLE participants also raises the possibility of pre-surgical functional re-organisation, such that semantic cognition has shifted away from the ATLs, in contrast to in SD and healthy controls where the semantic degradation occurs in the context of a previously intact system. However, there is evidence from direct cortical grid electrode studies that the bilateral ATLs remain as core semantic regions in TLE patients who require unilateral resection. Indeed, the maximal site of activation in the ventral ATL for semantic processing in these patients is identical to the peak site of semantic-related activation found in fMRI studies of healthy participants (32, 33).

## Results

### Demographic and clinical information

Table 1 displays demographic and clinical information for participants. Groups were matched for sex and years of education. In keeping with the inherent aetiological differences, both TLE groups were significantly younger on average than the neuropsychology controls and SD (*P* < 0.0001).

**Table 1.**
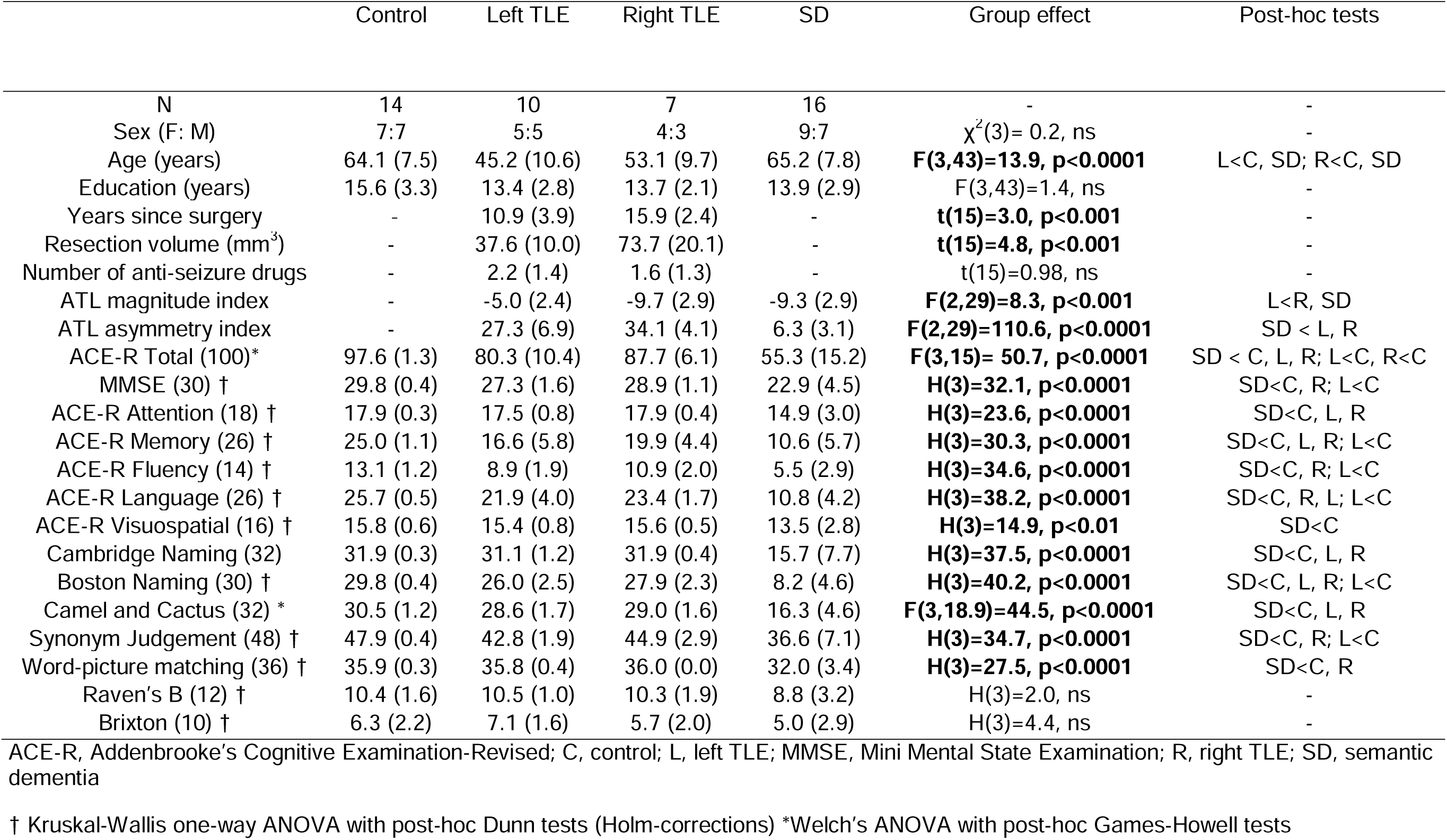
Demographic, clinical, imaging and neuropsychology results.

|  | Control | Left TLE | Right TLE | SD | Group effect | Post-hoc tests |
| --- | --- | --- | --- | --- | --- | --- |
| N | 14 | 10 | 7 | 16 | - | - |
| Sex (F: M) | 7:7 | 5:5 | 4:3 | 9:7 | $\chi^2(3) = 0.2$, ns | - |
| Age (years) | 64.1 (7.5) | 45.2 (10.6) | 53.1 (9.7) | 65.2 (7.8) | **F(3,43)=13.9, p<0.0001** | L<C, SD; R<C, SD |
| Education (years) | 15.6 (3.3) | 13.4 (2.8) | 13.7 (2.1) | 13.9 (2.9) | F(3,43)=1.4, ns | - |
| Years since surgery | - | 10.9 (3.9) | 15.9 (2.4) | - | **t(15)=3.0, p<0.001** | - |
| Resection volume (mm<sup>3</sup>) | - | 37.6 (10.0) | 73.7 (20.1) | - | **t(15)=4.8, p<0.001** | - |
| Number of anti-seizure drugs | - | 2.2 (1.4) | 1.6 (1.3) | - | t(15)=0.98, ns | - |
| ATL magnitude index | - | -5.0 (2.4) | -9.7 (2.9) | -9.3 (2.9) | **F(2,29)=8.3, p<0.001** | L<R, SD |
| ATL asymmetry index | - | 27.3 (6.9) | 34.1 (4.1) | 6.3 (3.1) | **F(2,29)=110.6, p<0.0001** | SD < L, R |
| ACE-R Total (100)* | 97.6 (1.3) | 80.3 (10.4) | 87.7 (6.1) | 55.3 (15.2) | **F(3,15)= 50.7, p<0.0001** | SD < C, L, R; L<C, R<C |
| MMSE (30) † | 29.8 (0.4) | 27.3 (1.6) | 28.9 (1.1) | 22.9 (4.5) | **H(3)=32.1, p<0.0001** | SD<C, R; L<C |
| ACE-R Attention (18) † | 17.9 (0.3) | 17.5 (0.8) | 17.9 (0.4) | 14.9 (3.0) | **H(3)=23.6, p<0.0001** | SD<C, L, R |
| ACE-R Memory (26) † | 25.0 (1.1) | 16.6 (5.8) | 19.9 (4.4) | 10.6 (5.7) | **H(3)=30.3, p<0.0001** | SD<C, L, R; L<C |
| ACE-R Fluency (14) † | 13.1 (1.2) | 8.9 (1.9) | 10.9 (2.0) | 5.5 (2.9) | **H(3)=34.6, p<0.0001** | SD<C, R; L<C |
| ACE-R Language (26) † | 25.7 (0.5) | 21.9 (4.0) | 23.4 (1.7) | 10.8 (4.2) | **H(3)=38.2, p<0.0001** | SD<C, R, L; L<C |
| ACE-R Visuospatial (16) † | 15.8 (0.6) | 15.4 (0.8) | 15.6 (0.5) | 13.5 (2.8) | **H(3)=14.9, p<0.01** | SD<C |
| Cambridge Naming (32) | 31.9 (0.3) | 31.1 (1.2) | 31.9 (0.4) | 15.7 (7.7) | **H(3)=37.5, p<0.0001** | SD<C, L, R |
| Boston Naming (30) † | 29.8 (0.4) | 26.0 (2.5) | 27.9 (2.3) | 8.2 (4.6) | **H(3)=40.2, p<0.0001** | SD<C, L, R; L<C |
| Camel and Cactus (32) * | 30.5 (1.2) | 28.6 (1.7) | 29.0 (1.6) | 16.3 (4.6) | **F(3,18.9)=44.5, p<0.0001** | SD<C, L, R |
| Synonym Judgement (48) † | 47.9 (0.4) | 42.8 (1.9) | 44.9 (2.9) | 36.6 (7.1) | **H(3)=34.7, p<0.0001** | SD<C, R; L<C |
| Word-picture matching (36) † | 35.9 (0.3) | 35.8 (0.4) | 36.0 (0.0) | 32.0 (3.4) | **H(3)=27.5, p<0.0001** | SD<C, R |
| Raven's B (12) † | 10.4 (1.6) | 10.5 (1.0) | 10.3 (1.9) | 8.8 (3.2) | H(3)=2.0, ns | - |
| Brixton (10) † | 6.3 (2.2) | 7.1 (1.6) | 5.7 (2.0) | 5.0 (2.9) | H(3)=4.4, ns | - |

ACE-R, Addenbrooke's Cognitive Examination-Revised; C, control; L, left TLE; MMSE, Mini Mental State Examination; R, right TLE; SD, semantic dementia
† Kruskal-Wallis one-way ANOVA with post-hoc Dunn tests (Holm-corrections) \*Welch's ANOVA with post-hoc Games-Howell tests

### Magnetic Resonance Imaging

Voxel-based morphometry was conducted to determine the location and extent of grey matter volume reduction in each patient group relative to age-matched controls. As expected, the SD group had significantly reduced grey matter in the bilateral ATLs. In contrast, each TLE group had one cluster of volume loss, in either the left or right ATL depending on the site of the neurosurgery (Fig. 1*A* and Table S1). Individual ATL indices were calculated to directly compare SD and TLE groups on total ATL volume loss and asymmetry of ATL volume loss. Magnitude and asymmetry indices are displayed in Fig. 1*B*. There were overlapping levels of ATL magnitude between SD and TLE; the SD and right TLE patients were matched on ATL magnitude (*P* = 0.96), whereas the left TLE cases had higher magnitude indices (i.e., greater ATL volume) than both SD (*P* < 0.01) and right TLE (*P* < 0.01) (Table 1). The difference in ATL magnitude between left and right TLE is in keeping with current surgical standards, where left ATL resections are more conservative to avoid disruption to language networks (88). Despite similar levels of ATL magnitude, there was a large difference in ATL asymmetry between SD and TLE (*F*(2, 29) = 110.57, *P* < 0.0001). Although most of the SD group were asymmetric to a degree (with most having left > right damage), this was far lower than in TLE, highlighting the bilateral atrophy in SD. Significant differences in ATL asymmetry were found between SD and left TLE (*P* < 0.0001) and between SD and right TLE (*P* < 0.0001). Although each TLE group had high levels of asymmetry, the right TLE group was more asymmetric on average than left TLE (*P* < 0.05) reflecting the larger resection volumes.

**Fig. 1.**
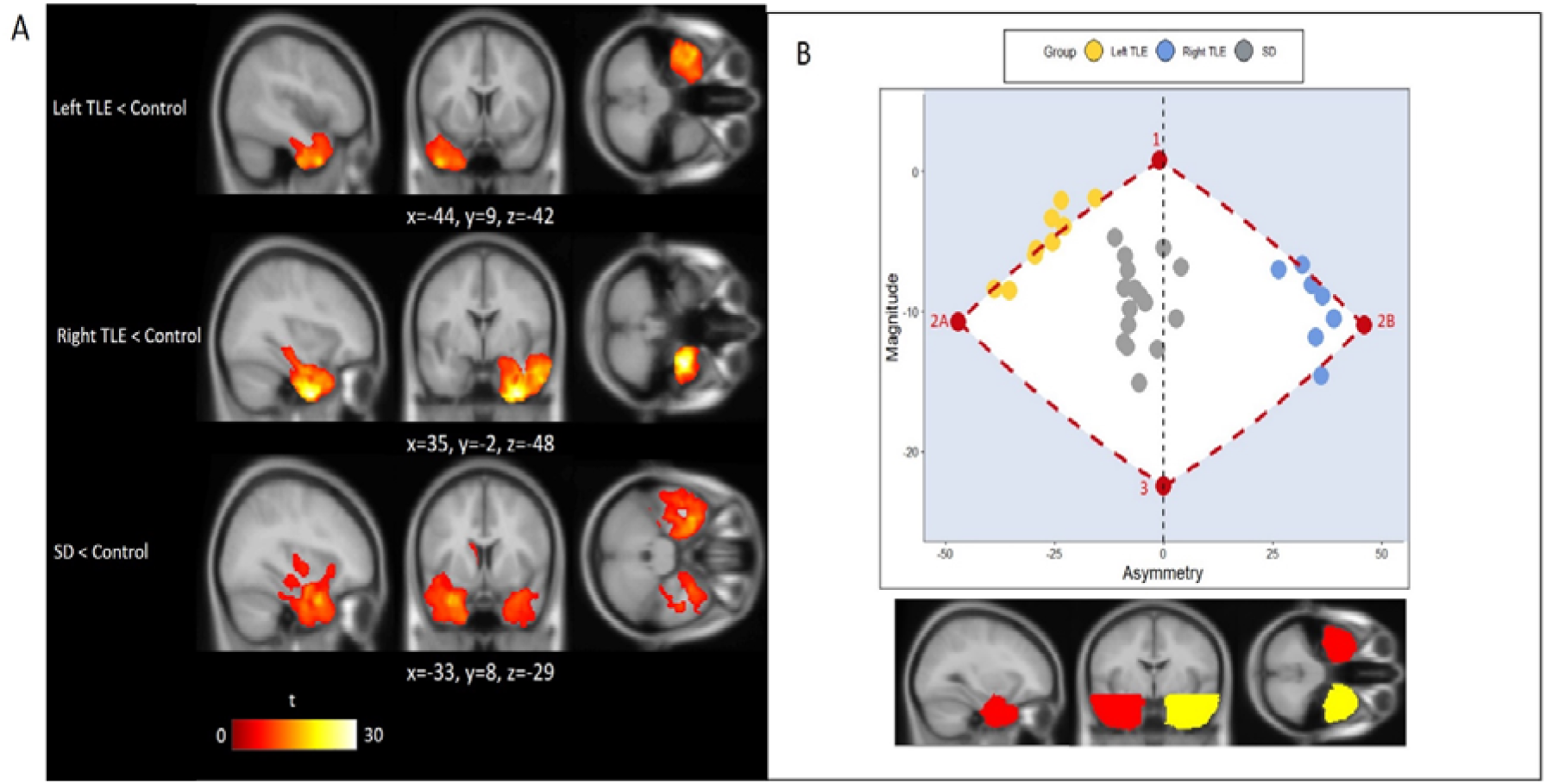
Neuroimaging results. (*A*) Voxel-based morphometry results showing clusters of reduced grey matter volume in (i) left TLE, (ii) right TLE and (iii) SD compared to controls. Images are thresholded using a threshold of *P*(FWE) < 0.05 with a cluster threshold of 100 voxels. Significant clusters are overlaid on the MNI avg152 T1 template. Co-ordinates are reported in Montreal Neurological Institute space. (*B*) Scatter plot displaying ATL magnitude and asymmetry indices for each patient. Lower magnitude values indicate greater volume loss, and negative asymmetry values indicate left>right ATL volume loss. The red points represent a marker showing from (i) 1-2A/B, the extremity boundary after purely unilateral left/right resection; and (ii) 2A/B-3 being the most extreme one could be with additional levels of bilateral damage, until (iii) 3 - complete bilateral resection. The anatomical location of the left and right ATL masks used for deriving the magnitude and asymmetry indices is displayed below the scatter plot. SD, semantic dementia; TLE, temporal lobe epilepsy.

### Semantic memory

Despite comparable volumes of overall ATL damage, the SD patients (bilateral damage) had considerably worse scores across the full range of semantic tasks than either TLE group or age-matched controls (Table 1). Generally, the left and right TLE groups were mildly impaired, with no left vs. right differences. The comparisons between TLE and controls that reached statistical significance after correcting for multiple comparisons were between left TLE and controls on the Boston Naming task (*P* < 0.05), Camel and Cactus (*P* < 0.05) and synonym judgement task (*P* < 0.01). In addition, the majority of the TLE sample (both left and right) (70.6%) had a semantic composite score below the control-derived lower bound of normality (Fig. 2), consistent with the presence of a mild, global semantic impairment. Full details of the post-hoc pairwise comparisons are reported in Table S2.

**Fig. 2.**
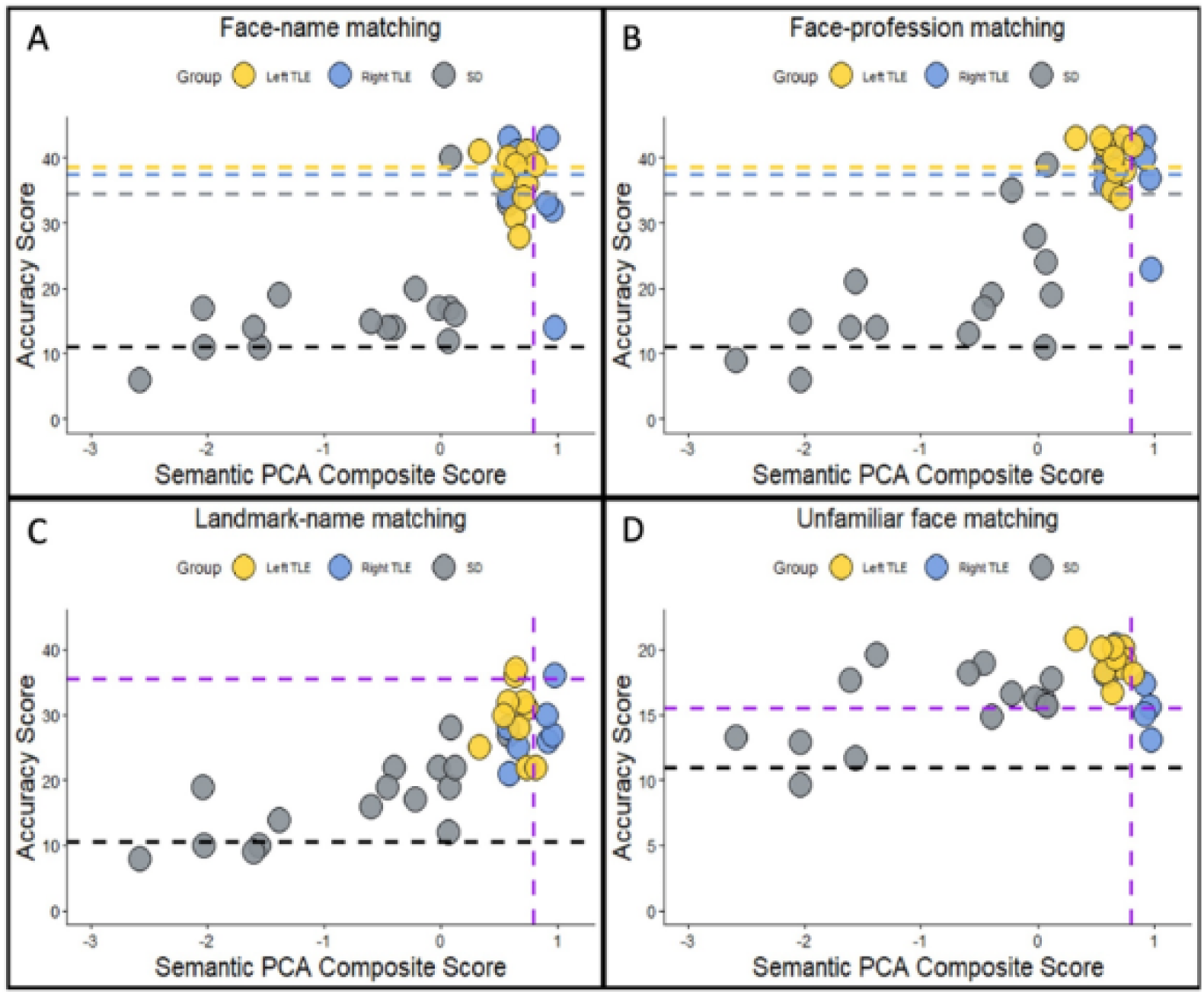
Neuropsychological performance plotted against semantic PCA composite scores for (*A*) face-name matching, (*B*) face-profession matching, (*C*) landmark-name matching and (*D*) unfamiliar perceptual face matching. The black horizontal line displays chance-level performance. The purple vertical line displays the control lower bound of normality (1.96 standard deviations below the control average). For (*A*) and (*B*) the yellow, blue and grey horizontal lines display the minimum score required in each group to not be impaired if they were the average age of their group (line colour corresponds to point colour). For (C) and (D) the purple horizontal line displays the score required to not be impaired (the same across all groups). PCA, principal component analysis; SD, semantic dementia; TLE, temporal lobe epilepsy.

### Person knowledge

In addition to impaired general semantic processing, the SD group displayed a simultaneous degradation of person knowledge. The same pattern was found after unilateral damage, where both TLE groups were impaired on person knowledge, although this was far milder than in SD. There was a significant main effect of group in both the face-name matching (*F*(3, 43) = 14.67, *P* < 0.001) and face-profession matching tasks (F(3, 43) = 15.99, *P* < 0.001). Controls performed better than left TLE (*P* < 0.01), right TLE (*P* = 0.07) and SD (*P* < 0.001) on the face-name matching task. A similar pattern was found in face-profession matching; SD (*P* < 0.001), left TLE (*P* < 0.01) and right TLE (*P* < 0.05) groups had poorer scores than controls, and SD were also worse than left TLE (*P* < 0.05).

All three patient groups were impaired on the landmark-to-name matching task, demonstrating that the person knowledge deficits found were not selective but generalised to another type of specific-level concept. There was a significant group effect on landmark-name matching (*F*(3, 16.7) = 63.7, *P* < 0.0001). Post-hoc Games-Howell tests revealed that each patient group performed worse than controls (*P* < 0.05), and SD performed worse than left and right TLE (*P* < 0.05).

Fig. 2 shows performance on each task plotted against semantic composite score for each individual patient. Across each group, most patients were impaired on the face-name matching (percentage impaired; left TLE = 70%, right TLE = 57.1%, SD = 93,8%) and on the difficulty-matched landmark-name matching task (percentage impaired; left TLE = 80%, right TLE = 85.7%, SD = 100%). Fewer patients were impaired on the face-profession matching task (percentage impaired; left TLE = 30%, right TLE = 42.9%, SD = 87.5%). As with general semantic memory, there were no left/right differences. No significant differences between left TLE and right TLE were found for face-name matching (*P* = 0.95), face-profession matching (*P* = 0.99), or landmark-name matching (*P* = 0.85).

### Perceptual Face matching

#### Accuracy

To explore the impact of bilateral vs. unilateral (left vs. right) ATL damage on perceptual processes, we first examined face matching performance accuracy on the unfamiliar condition only. There was a significant main effect of group on unfamiliar face matching accuracy (*F*(3, 43) = 6.82, *P* < 0.001), due to poorer performance by the SD group than both controls (*P* < 0.01) and left TLE (*P* < 0.01). This effect, however, was driven by a minority of severely impaired SD patients who had the greatest degree of overall semantic impairment (Fig. 2*D*). Surprisingly, a few of the right TLE patients were impaired on this task (percentage impaired; left TLE=0%, right TLE=28.6%, SD = 31.3%).

Next, we further explored the contribution of the ATLs to perception by comparing perceptual face matching performance for famous vs. unfamiliar faces. A familiarity effect was found for all groups - a mixed ANOVA yielded significant main effects of group (*F*(3, 43) = 6.75, *P* < 0.001) and face stimulus type (*F*(3, 43) = 37.78, *P* < 0.0001) (Fig. 3*A*). Each group was significantly more accurate for matching famous faces compared to unfamiliar faces (controls, *P* < 0.001; left TLE, *P* = 0.06; right TLE, *P* < 0.05; SD, *P* < 0.001).

**Fig. 3.**
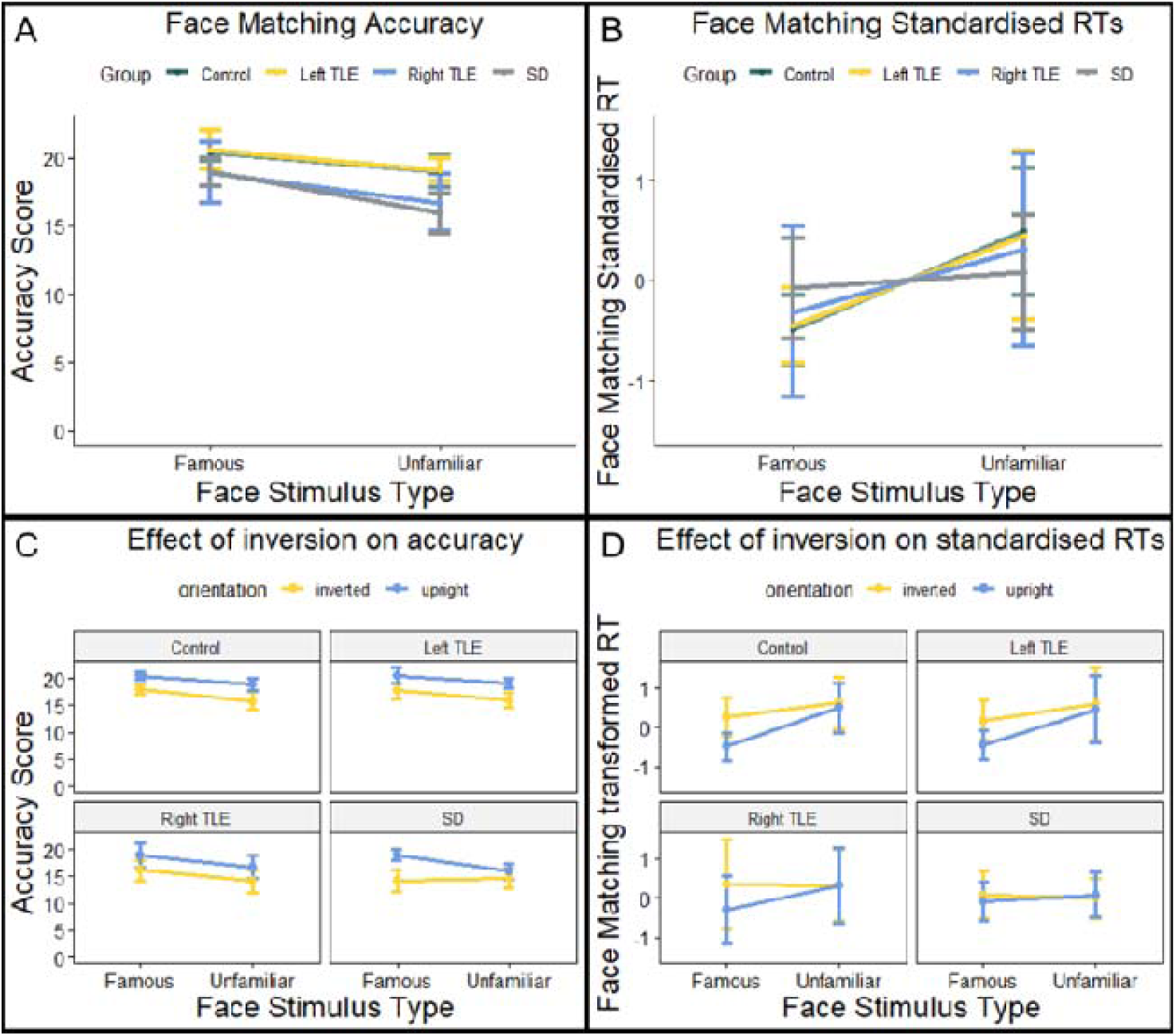
Mean accuracy and standardised RTs on the perceptual face matching task. (*A*) Face matching accuracy scores for famous vs. unfamiliar faces. (*B*) Face matching standardised RTs for famous vs. unfamiliar faces. (*C*) Face matching accuracy scores for famous vs. unfamiliar faces in both upright and inverted conditions. (*D*) Face matching standardised RTs for famous vs. unfamiliar faces in both upright and inverted conditions. Bars display 95% confidence intervals. SD, semantic dementia; TLE, temporal lobe epilepsy.

#### Response times

As a further assessment of a potential contribution of the ATL to perception, RTs of correct responses to famous vs. unfamiliar faces were compared. To account for differences in baseline RTs between groups, a z-score transformation was applied to the raw RTs (89). Raw RTs (Fig. S1) were standardised for each participant by taking the RT for each familiarity condition, subtracting the group mean RT (across both conditions) and dividing by the standard deviation of the group RT. This method has been used previously to account for slower baseline responding in SD (90). All groups produced faster responses to famous than to unfamiliar faces, although the effect was reduced in SD and right TLE. A mixed ANOVA revealed an interaction between group and familiarity (*F*(3, 43) = 3.98, *P* < 0.05) (Fig. 3*B*). Post-hoc tests revealed that although RTs for famous faces were faster across each group, this difference only reached significance in controls and left TLE (controls, *t* = 4.83, *P* < 0.001; left TLE, *t* = 3.63, *P* < 0.01, right TLE, *t* = 1.53, *P* =0.18; SD, *t* = 1.95, *P* = 0.07).

#### RTs across different levels of person knowledge

The correspondence between item-specific semantic status and perceptual performance was assessed by categorising face matching trials into ‘fully known’, ‘partially known’ or ‘unknown’, based on semantic performance in the person knowledge tasks. This method allows perceptual performance to be compared across items and has been used in previous studies of visual recognition in SD (91, 92). As there were very few ‘unknown’ trials in the controls and TLE groups, the ‘unknown’ and ‘partially known’ trials were combined into a single category. Similarly, ‘fully known’ and ‘partially known’ were combined in the SD group due to a lack of ‘fully known’ trials. In all groups, face matching RTs were quicker for ‘fully known’ trials (i.e. with the most semantic knowledge), further highlighting the association between semantic knowledge and perception (Fig. 4) There was a main effect of semantic knowledge in controls (χ^2^(2) = 19.8, *P* < 0.0001), with faster RTs for ‘fully known’ items compared to ‘partially known’/’unknown’ (*P* < 0.001) and unfamiliar items (*P* < 0.001). There was also a main effect of semantic knowledge in left TLE (χ^2^(2) = 12.6, *P* < 0.01) with faster RTs for ‘fully known’ than ‘partially known’/’unknown’ (*P* < 0.05) and unfamiliar items (*P* < 0.01). There was a significant main effect in the SD group also (χ^2^(2) = 6.93, *P* < 0.05), with faster RTs for ‘fully/partially known’ compared to unfamiliar items (*P* < 0.05). Surprisingly, there was no significant effect of semantic knowledge in right TLE (χ^2^(2) = 2.57, *P* = 0.28).

**Fig. 4.**
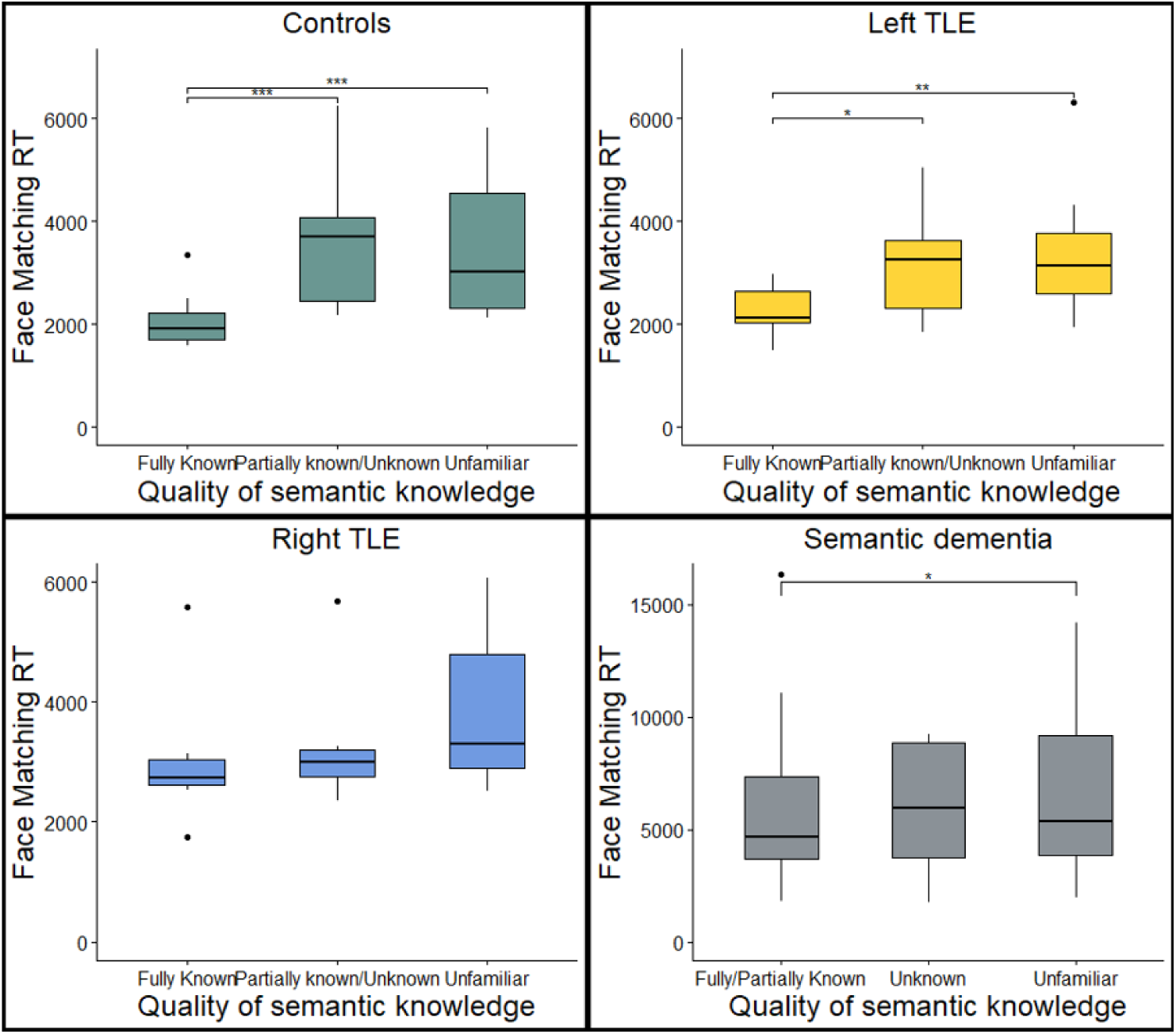
Perceptual face matching RTs for different levels of semantic knowledge. (*A*) controls, (*B*) left TLE, (*C*) right TLE and (*D*) semantic dementia. TLE; temporal lobe epilepsy. \**P* < 0.05, \*\**P* < 0.01, \*\*\**P* < 0.001

#### The effect of inversion on perceptual face matching

To explore whether the familiarity effect persisted even when faces were inverted, three-way mixed ANOVAs were conducted on face matching accuracy and standardised RTs separately, with group, face stimulus type and face orientation as factors. There was a significant three-way interaction between these factors on face matching accuracy (*F*(3, 43) = 4.06, *P* < 0.05) (Fig. 3*C*). Inversion abolished the familiarity effect on accuracy in SD but not in patients with unilateral damage or in healthy controls. Separate ANOVAs on each group revealed a two-way interaction between face stimulus type and orientation in SD (*F*(1, 15) = 9.54, *P* < 0.01), but not in controls (*F*(1, 13) = 0.56, *P* = 0.47), left TLE (*F*(1, 9) = 0.04, *P* = 0.84) or right TLE (*F*(1, 6) = 0.03, *P* = 0.87). There were significant two-way interactions between group and face stimulus type (*F*(3, 43) = 5.82, *P* < 0.01), group and face orientation (*F*(3, 43) = 11.04, *P* < 0.0001), and face stimulus type and face orientation (*F*(3, 43) = 18.02, *P* < 0.001) (Fig 3*D*). There was no three-way interaction between group, face stimulus type and face orientation on standardised face matching RTs (*F*(3, 43) = 0.55, *P* = 0.65).

## Discussion

This study clarifies the function of the ATL with relevance to cognitive and clinical neuroscience. Parallel theories and literatures have been developed, each supported by findings from neuropsychology, functional neuroimaging and other techniques. The discovery that the ATLs respond to familiar faces and that ATL damage causes face recognition deficits has led to proposals that the ATLs should be considered part of an extended face processing network (1–4). A separate substantial literature proposes that the ATLs are crucial to the formation and activation of all concepts and at all ‘levels’ from superordinate (e.g., animals or objects), to intermediate (e.g., humans or buildings) to specific (e.g., Marilyn Monroe, the Eiffel Tower), with increasing demands on ATL processing along this succession of levels. The global aim of this investigation was to bring these two parallel literatures together, via both general semantic and face processing tasks applied to patients with two different types of ATL damage: bilateral atrophy in semantic dementia vs. unilateral resection in treatment for epilepsy.

Although most of the SD cases had, as is typical, a degree of ATL asymmetry, the distribution of volume loss was clearly bilateral in contrast to the unilateral loss following resection. Consequently, we were able to explore the impact of bilateral vs. unilateral (left or right) ATL damage on (a) general semantic memory, (b) person knowledge, and (c) the perceptual processes that are primarily considered to be functions of ventral occipitotemporal areas. In the following sections we integrate the key findings of the study within a unified neurocognitive framework for the ATLs in face recognition, person knowledge and semantic memory, and discuss the implications for the extended face network (1–3).

### 1 The ATLs support a singular, common semantic system

Bilateral ATL damage generated substantial impairments in both general semantic processing and person knowledge. The semantic impairment occurred for all types of stimuli and across expressive and receptive tasks, in line with the global degradation of conceptual knowledge characteristic of SD (29, 70). This study highlighted the consequences of bilateral ATL damage on person knowledge, supporting previous neuropsychological investigations in SD (10, 34). Unilateral ATL damage also caused dual impairments in general semantic memory and person knowledge, although to a much milder degree than the bilateral damage in SD. This finding mirrors previous studies of ATL-resected patients, which have reported a subtle generalised semantic impairment following resection of either the left or right ATL (67, 68, 93). Taken together, the results from the two patient groups are consistent with a semantic system underpinned by the bilateral ATLs that represents all types of conceptual knowledge, including person knowledge (5, 6).

Knowledge of famous people was severely impaired by bilateral ATL damage, and many SD patients performed around chance-level on the tasks assessing this cognitive sphere. Bilateral damage caused a similarly severe deficit in a landmark knowledge task, which was included as it taps into another type of specific-level concept/‘unique entity’ exemplar (94, 95). Neuropsychological studies have demonstrated that the semantic decline in SD is graded, such that specific-level individuations (e.g. differentiating between a dalmatian and other breeds of dog) are more vulnerable than more basic semantic distinctions (e.g. differentiating between a dalmatian and other types of mammal) (96–99). Consequently, tasks requiring specific-level distinctions are the most sensitive assessments of semantic integrity (99). Clear impairments for the specific-level concepts were also found after unilateral damage (although much milder than in SD), in line with previous findings that the semantic deficits from unilateral ATL damage are amplified when more challenging tasks or concepts are used (67, 68, 100). The results here indicate therefore that, although all concepts are supported by the ATLs, the representations in the semantic system are such that specific-level concepts (of which individual people or landmarks are examples) are inherently more vulnerable to mild damage (as simulated in multiple implemented computational instantiations of the hub-and-spoke model) (101, 102).

### 2 The functionally-unitary semantic system is supported bilaterally

There were no selective semantic deficits after either left or right unilateral ATL damage: both were characterized by a mild generalised semantic impairment. This finding implicates the bilateral ATLs as important for conceptual knowledge, a proposal which is supported by convergent evidence from studies in patients and in healthy participants. FMRI studies consistently detect bilateral ventrolateral ATL activation when healthy participants engage in semantic processing (provided appropriate techniques are used to maximise the otherwise ‘shy’ ventral ATL signal) (30, 103). Furthermore, local field potentials in overlapping bilateral ventrolateral ATL regions have been detected from grid electrode recordings during semantic tasks in pre-resected patients (32). Causal evidence for the ATLs in semantic memory has also been gained from neurostimulation studies: both TMS to either the left or right ATL in healthy participants (32) and direct cortical stimulation of the left or right ventrolateral ATL (33) produce a transient slowing of semantic processing but not non-semantic processing.(31) There is evidence that functional connectivity between the ATLs increases during challenging semantic tasks in healthy participants (104) with the degree of functional connectivity predicting semantic performance (104) as well as behavioural outcome after stroke (105). Consequently, it appears that the ATLs work together as a single semantic system, where both the (i) integrity of the left and right ATL and the (ii) functional connectivity between the ATLs are crucial.

Despite similarities in the *quality* of the semantic impairment, there were differences in the *magnitude* of the impairment from bilateral vs. unilateral ATL damage. The finding of mild impairment after unilateral damage vs. severe deficits after bilateral damage mirrors previous neuropsychological investigations (67, 70) and also fits with the classical comparative neurology literature, where bilateral ATL ablation in macaques (and in one human case) generates a severe multimodal associative agnosia, yet unilateral resection yields only a mild and transient effect (106, 107). Strikingly, there was considerable overlap in total ATL damage across the two groups, meaning that, whilst the level of semantic impairment is governed by the overall level of ATL damage (21, 84), the uni-/bi-lateral distribution of damage is also crucial.

One advantage of a bilaterally implemented semantic system is the inclusion of partial redundancy, which would configure the semantic system to be robust to unilateral damage. This hypothesis has been captured by formal computational models, which have demonstrated that when a semantic hub was divided into ‘demi-hubs’ (mimicking the left and right ATLs), simulated unilateral damage generated a much milder impairment than bilateral damage, even when total damage was kept constant (59). After unilateral damage, distorted semantic representations were restricted to one demi-hub, and the propagation of activation noise was constrained to ipsilateral units. As a result, the undamaged contralateral demi-hub was able to function with relative accuracy albeit more slowly (59). Insights into the compensatory neural mechanisms underlying the ATL’s resilience to unilateral damage/perturbation have been derived from fMRI studies, where, after unilateral damage/perturbation (either from resection or rTMS), the unaffected contralateral ATL not only upregulates its activity but increases its effective connectivity with the affected ATL (60, 81, 108).

### 3 The ATL-semantic system interacts with posterior temporal regions to support face perception

Although the ATL is critical for semantic memory, there was no evidence that this region is similarly critical for the ability to discriminate between faces based on visual properties. Face perception abilities were preserved after either bilateral or unilateral ATL damage (except for a minority of severe SD patients). This result aligns with previous findings of intact face perception abilities alongside a preservation of perceptual skills broadly in SD (8, 20, 25, 109).

The contribution of ATL-based semantics to face perception was explored by assessing the classic face familiarity effect. The familiarity effect was robustly replicated in healthy participants, even when faces were inverted (although, as expected in this classic paradigm, to a lesser degree than upright). In contrast to the patients’ preserved face-matching abilities, bilateral ATL damage and associated semantic degradation diminished the familiarity effect. The relationship between semantic knowledge and face perception was further highlighted by the finding of item-specific correspondence between the quality of semantic knowledge and the strength of the familiarity effect, across all participant groups. Inversion completely obliterated the familiarity effect in bilateral ATL cases, which implies that the semantic contribution to face perception is maintained when faces are inverted but is more subtle and thus more sensitive to semantic degradation.

One potential explanation of the familiarity effect is that it reflects interactivity between the ATL and ventral occipitotemporal cortex. During perception of famous faces, the activated semantic system feeds back expectations/predictions about the input to support the early stages of visual processing. Bilateral ATL damage would result in degraded and diminished semantic representations being projected back to posterior perceptual areas, thus disrupting any facilitation or acceleration that is provided by a healthy semantic system. This proposal can be accommodated within the hub-and-spoke model of semantic memory where, through its interactivity and connectivity, the ATL-semantic hub not only receives inputs from modality-specific posterior areas but also projects back to them (102). Depth electrode recordings have detected initial ‘first-pass’ activation in the ATLs during visual recognition, which then may feedback activated semantics to posterior temporal cortex (110). In addition, there is electrophysiological evidence that semantic information modulates ERPs associated with early visual processing (111–114). Further evidence for an interaction between conceptual and perceptual systems derives from people with SD who are impaired on perceptual tasks such as object recognition (47), word recognition (90) and object/lexical decision (48) with the perceptual impairment aligning with the level of semantic degradation. Most strikingly, when SD patients are asked to copy line drawings of real objects/animals a mere 10 seconds after the stimulus pictures have been withdrawn, their degraded semantic systems delete item-specific features (e.g., a camel’s hump) and include properties that are true more generally of that class but not of the specific concept just presented (e.g., drawing a duck with four legs) (115).

### 4 Graded functional differences between the ATLs emerge through different connectivity strengths with modality-specific regions

Although there were no significant differences in semantic performance after left vs. right ATL resection, people with unilateral right ATL damage performed more poorly at perceptual face matching than their left-sided counterparts, in terms of reduced accuracy and a diminished familiarity effect. Face recognition problems have previously been reported after right ATL damage from unilateral resection for TLE (22, 24, 56, 68) and also right>left ATL atrophy in semantic dementia (23, 116). According to the hub-and-spoke model, although conceptual knowledge is represented bilaterally, graded asymmetries may emerge from different connectivity strengths of the left and right ATL with modality-specific regions. As a result, although all aspects of semantic memory would be impaired by ATL damage, some types of semantic category or task may be disproportionately affected if the damage is asymmetric (5, 61, 62). The most reliable example is anomia, which is more severe after left ATL damage in both semantic dementia (62, 63) and unilateral resection for TLE (22, 68). The increased anomia from left ATL damage has been attributed to the region having stronger connections with left-lateralised speech production areas, a proposal which has been captured computationally (63).

There is a right-sided dominance for face processing in the posterior ventral temporal cortex (51, 54, 64). In the posterior ventral temporal cortex, there are graded asymmetries in functional organisation rather than absolute differences between the hemispheres, such that face processing is supported bilaterally but more strongly in the right hemisphere (51, 117). The increased face recognition problems after right sided ATL damage might reflect downstream effects of this functional asymmetry, i.e., the stronger visual input from the right posterior temporal cortex is projected to the right ATL (64).

Relative specialisations *within* the ATLs may also emerge via the same principle of graded connectivity (5, 61, 118, 119). For example, there is fMRI evidence that, in addition to activation in a core ventrolateral ATL ‘hotspot’, person knowledge (faces and written names of famous people) elicits weaker yet selective activation in a slightly anterior ATL subregion (36). The temporal poles are most strongly connected to the orbitofrontal cortex via the uncinate fasciculus (120) leading to speculation that the relative preference of this ATL subregion for person knowledge reflects its proximity to paralimbic regions, which may represent ‘spokes’ particularly important for the formation of person knowledge (36, 121).

### Implications for the extended face network

Our findings have three key implications for the extended face network. First, the core function of the ATL in face recognition is the representation of semantic memory. Damage to the ATL does not impair the perceptual processes necessary for face perception, which instead depend on ‘core’ face recognition areas in more posterior temporal regions (37, 38). Rather, ATL damage degrades the semantic representations which are needed to support familiar face recognition through the provision of activated semantics. Critically, the ATLs are not face-selective, but support person knowledge as part of a transmodal semantic representational system.

Secondly, the extended network is interactive in nature. Rather than a purely feedforward hierarchical ventral pathway, the core posterior temporal face perception areas interact bidirectionally with ATL-semantic regions (that code information about people – not just faces, alongside all other concepts). Accordingly, activated semantics project back expectations/predictions about the input to support the early stages of visual processing, via rapid feedback along the inferior longitudinal fasciculus. Following this semantic feedback from the ATL, perceptual demands are reduced when faces are familiar, which leads to a boost or acceleration of recognition.

Thirdly, the extended face network recruits the ATLs bilaterally. Existing models of face recognition have not made strong claims on the differential roles of the left/right ATL in the extended face recognition network (1–3), although the nature of the discussion about core areas of the face recognition network is itself predominantly “right-lateralised” (51–54). In this study we demonstrated that person knowledge is supported by the left and right ATL, as part of a broader conceptual representational system. However, the right ATL may be relatively more important for face recognition because it receives increased visual input from right posterior temporal ventral cortex.

## Data Sharing Plans

Due to the limits of the ethics approval for these patient studies, the data cannot be openly shared. Requests for suitably anonymised data can be addressed to the senior author and may require a data transfer agreement.

## Acknowledgments

We thank the patients and their families or caregivers for giving up the time to take part in the study. We also thank Dr Thomas Cope, Dr Sian Thompson and Dr Sofia Toniolo for their assistance with recruitment.

## Funding

M.A.R is supported by the Medical Research Council (SUAG/096 G116768). A.D.H is supported by the Medical Research Council (Career Development Award: MR/V031481/1). A.V was supported by the Université de Lorraine for this project (DrEAM/LUE grant). J.B.R is supported by the Medical Research Council (MC_UU_00030/14; MR/T033371,1), Wellcome Trust (220258), and the NIHR Cambridge Biomedical Research Centre (NIHR203312). M.A.L.R is supported by a Medical Research Council programme grant (MR/R023883/1) and intramural funding (MC_UU_00005/18). The views expressed are those of the authors and not necessarily those of the NIHR or the Department of Health and Social Care.

## Notes

### Competing Interest Statement

The authors have declared no competing interest.

### Author Declarations

The National Research Ethics Service's East of England Cambridge Central Committee gave ethical approval for this work(IRAS project ID: 252986)

