## Supplementary Materials for "The impact of bilateral versus unilateral anterior temporal lobe damage on face recognition, person knowledge and semantic memory"

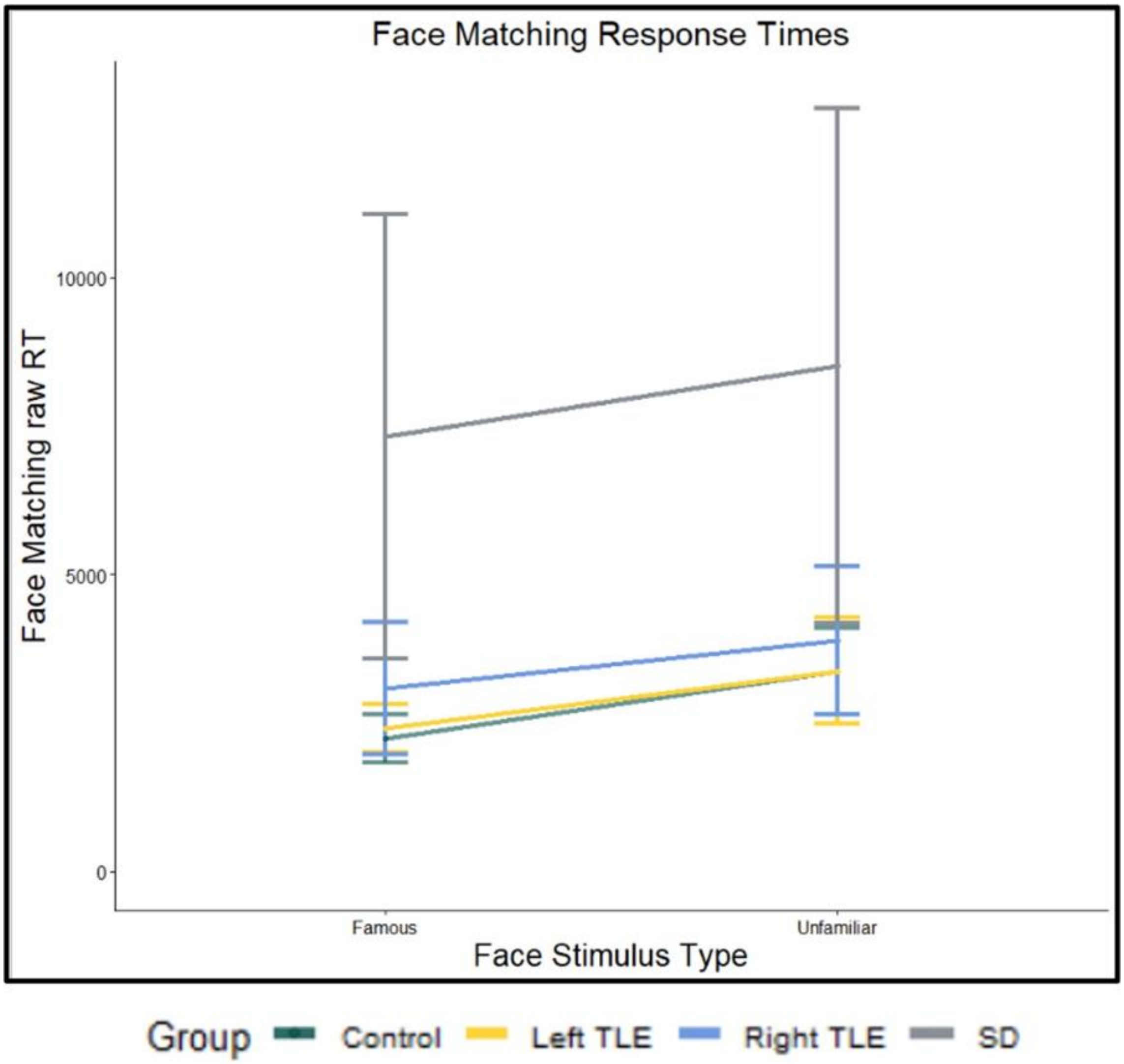


**Fig. S1.** Raw RTs on the perceptual face matching task. Bars display 95% confidence intervals.

**Table S1.** Voxel based morphometry results showing regions of reduced grey matter volume in each patient group relative to controls

| Regions | Hemisphere | Number of voxels | Peak MNI co-ordinate | | | Peak MNI co-ordinate region | *t*-value |
|---|---|---|---|---|---|---|---|
| | | | *x* | *y* | *z* | | |
| *Left TLE < Controls* | | | | | | | |
| | Left | 12,093 | -44 | 9 | -42 | Temporal pole | 18.50 |
| *Right TLE < Controls* | | | | | | | |
| | Right | 16,772 | 35 | -2 | -48 | Inferior temporal gyrus | 29.78 |
| *SD < Controls* | | | | | | | |
| | Left | 21,630 | -33 | 8 | -29 | Temporal pole | 16.98 |
| | Right | 6,810 | 33 | 6 | -27 | Temporal pole | 11.77 |
| | Left | 685 | -53 | -53 | 3 | Middle temporal gyrus | 8.88 |

MNI, Montreal Neurological Institute; SD, semantic dementia; TLE, temporal lobe epilepsy

Clusters reported if significant at *P*(FWE) < 0.05 with a cluster threshold of 100 voxels.

**Table S2**. P-values for the post-hoc comparisons

| | C v L | C v R | C v SD | L v R | L v SD | R v SD |
|---|---|---|---|---|---|---|
| ACE-R total* | <0.01 | <0.05 | <0.0001 | ns | <0.001 | <0.0001 |
| MMSE† | <0.01 | ns | <0.0001 | ns | ns | <0.05 |
| ACE-R Attention† | ns | ns | <0.0001 | ns | <0.05 | <0.01 |
| ACE-R Memory† | <0.01 | ns | <0.0001 | ns | ns | <0.05 |
| ACE-R Fluency† | <0.01 | ns | <0.0001 | ns | ns | <0.01 |
| ACE-R Language† | <0.05 | ns | <0.0001 | ns | <0.05 | <0.05 |
| ACE-R Visuospatial† | ns | ns | <0.01 | ns | ns | ns |
| Cambridge Naming† | ns | ns | <0.0001 | ns | <0.001 | <0.001 |
| Boston Naming† | <0.05 | ns | <0.0001 | ns | <0.05 | <0.01 |
| Camel and Cactus* | <0.05 | ns | <0.0001 | ns | <0.000 | <0.0001 |
| Synonym Judgement† | <0.01 | ns | <0.0001 | ns | ns | <0.05 |
| Word to Picture Matching† | ns | <0.01 | <0.0001 | ns | <0.001 | ns |

ACE-R, Addenbrooke’s Cognitive Examination-Revised; C, control; L, left TLE; MMSE, Mini Mental State Examination; R, right TLE; SD, semantic dementia

* Games-Howell test
† Dunn test (Holm-corrections)
